# *GALC* variants affect glucocerebrosidase to galactosylceramidase activity ratio

**DOI:** 10.64898/2026.10.05.26364651

**Authors:** Amirhossein Rajabi Vajargah, Konstantin Senkevich, Zhao Zhang, Lang Liu, Saeid Amiri, Morvarid Ghamgosar Shahkhali, Meron Teferra, Camille Lacarriere-Keita, Pablo Sardi, Ming Sum Ruby Chiang, Cheryl Waters, Adil Harroud, Roy N. Alcalay, Thomas M Durcan, Edward A Fon, Ziv Gan-Or

**Affiliations:** Department of Neurology and Neurosurgery, McGill University, Montreal, QC, Canada; The Neuro (Montreal Neurological Institute-Hospital), Montreal, QC, Canada; Department of Human Genetics, McGill University, Montreal, QC, Canada; Quantitative Life Sciences, McGill University, Montreal, QC, Canada; Department of Specialized Medicine, Division of Medical Genetics, McGill University Health Centre, Montreal, Quebec, Canada; Department of Neurology, College of Physicians and Surgeons, Columbia University Medical Center, NY, USA; Gray Faculty of Medical & Health Sciences, Tel Aviv University, Tel Aviv, Israel; The Movement Disorders Institute, Department of Neurology, Sheba Medical Center, Tel Hashomer, Israel; Early Drug Discovery Unit (EDDU), Montreal Neurological Institute-Hospital (The Neuro), Montreal, QC, Canada; Rare and Neurologic Diseases Therapeutic Area, Sanofi, Framingham, MA 01701, USA; Victor Phillip Dahdaleh Institute of Genomic Medicine, McGill University, Montréal, QC, Canada; Center for Molecular Medicine, Department of Clinical Neuroscience, Karolinska Institutet, Stockholm, Sweden

**Author notes:** **Correspondence:** Ziv Gan-Or, The Neuro (Montreal Neurological Institute-Hospital) 1033 Pine Avenue, West, Ludmer Pavilion, room 312 Montreal, QC, H3A 1A1.

## Abstract

**Objective:** Genes within the lysosomal glycosphingolipid pathway, including *GBA1* and *GALC*, are important genetic risk factors in Parkinson’s disease (PD). We hypothesized that PD risk relates not only to individual enzyme effects but also to coordination between glucocerebrosidase (GCase) and galactosylceramidase (GalCase) activities, and examined how genetic variants influence their activity ratio.

**Methods:** We studied 1,290 individuals from the Parkinson’s Precision Medicine Initiative and Columbia University cohorts with genetic and dried-blood-spot enzymatic data. Genome-wide association analyses of the glucocerebrosidase/galactosylceramidase activity ratio were performed in the combined cohort, by sex, and per cohort, followed by meta-analysis. For lead variants, Pearson correlations between enzyme activities were compared across genotypes, and genotype-by-GCase interactions were tested.

**Results:** The *GALC* locus was associated with glucocerebrosidase/galactosylceramidase activity ratio in the combined cohort (rs365448; beta=-1.90; P=5.91e-115), replicated across cohorts and meta-analysis (beta = −1.66, P=6.33e-191), and in linkage disequilibrium with the PD GWAS index variant rs979812. This association remained significant in both sexes. Correlation between glucocerebrosidase and galactosylceramidase activities differed by rs979812 genotype (TT r=0.33 versus GG r=0.13; P=0.01), with a significant genotype-by-GCase interaction (P=3.05e-5). Among 134 PD loci examined, ten showed nominal association with the ratio-GWAS, including *GBA1*, where risk-variant carriers showed a similar pattern, yet not statistically significant.

**Interpretation:** *GALC* variants influence the balance between glucocerebrosidase and galactosylceramidase activity, and this relationship may differ by genotype at PD-associated variants. These findings suggest PD risk at lysosomal glycosphingolipid genes may partly reflect altered coordination between pathway enzymes rather than isolated single-enzyme effects, warranting functional validation.

## Introduction

Parkinson’s disease (PD) is a heterogeneous disorder caused by a combination of environmental exposures, genetic factors, and the natural process of aging. Several genetic subtypes of PD have been identified, for example, those associated with *GBA1* or *LRRK2* variants, likely involving distinct mechanisms.^1^ In addition, numerous common genetic risk factors for PD have been identified through genome-wide association studies (GWAS), with the most recent study reporting 159 independent associations across 134 loci.^2^ The mechanisms underlying the associations between the vast majority of these variants and PD remain unknown. For most loci, the specific genes and causal variants driving the associations have yet to be determined, despite ongoing computational and functional efforts to characterize them.^3, 4^

However, many genes implicated in PD converge on the lysosomal glycosphingolipid metabolism pathway, including *GBA1*,^5^ *GALC*,^6^ *SMPD1*,^7^ *ASAH1*,^8^ and others.^9^ Both common and rare variants in *GBA1* are reported in 5–30% of patients with PD,^10, 11^ including coding variants that likely affect the structure and/or function of glucocerebrosidase (the enzyme encoded by *GBA1*),^10^ as well as non-coding variants that likely influence glucocerebrosidase splicing and may also result in a dysfunctional enzyme.^12^ Galactosylceramidase, a lysosomal enzyme encoded by *GALC*, operates within the same lysosomal pathway as glucocerebrosidase. Both enzymes degrade glycosphingolipids, galactosylceramides and glucocerebrosides, respectively, and producing a monosaccharide (galactose or glucose) and ceramide.^13, 14^ *GALC* is the primary candidate gene within a GWAS locus on chromosome 14, as supported by both computational and functional studies. ^4, 6^ Previous work has shown that PD-associated variants in the *GALC* locus are linked to its expression and to the activity of galactosylceramidase. ^6^ Interestingly, a recent sex-stratified GWAS identified the *GALC* locus to be associated with PD only in females and not in males. ^15^

Given that multiple genes within the lysosomal glycosphingolipid metabolism pathway are implicated in PD, we hypothesize that disease risk is influenced not only by the effects of individual variants on specific gene functions, but also by the relationships among these enzymes. Specifically, how their activities correlate and collectively influence metabolic flux within the pathway. To test this hypothesis for *GBA1* and *GALC*, we designed a study to examine the relationship between the activities of their encoded enzymes using an activity ratio, and to assess how genetic variants influence this ratio.

## Methods

### Population

The study population consisted of two separate cohorts with available genetics data and enzymatic activity of glucocerebrosidase and galactosylceramidase from the Parkinson’s Progression Markers Initiative (PPMI)^16^ and Columbia University. ^6, 17^ Cohort demographics for the individuals with both genetics and enzymatic data available after quality controls consists of 360 individuals in PPMI and 930 individuals in Columbia cohort (N_total_ = 1290, N_females_ = 522, N_males_ = 768) with 427 controls and 863 PD cases. Diagnosis inclusion and exclusion criteria for Columbia^18^ and PPMI have been previously described (https://www.ppmi-info.org/study-design/study-cohorts).

### Genetic data

Genetic data for Columbia cohort consists of genotyping data which was performed with the OmniExpress GWAS array according to the manufacturer’s protocols (Illumina Inc.). Sample level and variant level quality control were performed as previously described (https://github.com/neurogenetics/GWAS-pipeline). In summary, samples that were heterozygosity outliers (inclusion criteria of −0.15 ≤F≤ 0.15), call rate outliers (missingness > 5%), had mismatched genetic and reported sex, or were identified as European ancestry outliers based on HapMap3 principal component analysis (PCA) in plink v1.9^19^ were removed. Additionally, we removed samples with relatedness closer than third degree relatives (pihat >0.125). Individual variants were excluded on the basis of variant missingness (> 5%), differences in missingness between cases and controls (p< 1e −04), haplotype missingness (p< 1e −04), and deviation from the Hardy–Weinberg equilibrium in controls (p< 1e −04). All participants were of European descent, confirmed by principal component analysis. Imputation was performed on filtered data with the Michigan Imputation Server using the Haplotype Reference Consortium reference panel r1.1 2016 and default settings Whole-genome sequencing data for the PPMI cohort were extracted from AMP-PD (v4 release; https://amp-pd.org/). Initial quality control was performed by AMP-PD as previously described (https://amp-pd.org/whole-genome-data). Additional quality control was applied, including filtering out variants with depth of coverage DP ≤ 20, removing samples with >5% missing genotypes, excluding individuals with excess heterozygosity (F < −0.15 or F > 0.15), and removing samples with call rate <95%. Individuals of non-European ancestry and those related closer than third-degree (pi-hat > 0.125) were also excluded.

After merging of the two cohorts, common variants with allele frequency > 0.05 were retained, individuals and variants with >5% missingness, and variants deviating from Hardy-Weinberg equilibrium in the control group (P < 1 × 10^-6^) were removed.

### Enzymatic activity

The enzymatic activities of glucocerebrosidase and galactosylceramidase in the Columbia cohort were measured from dried blood spot, as previously described.^20, 21^ In brief, enzyme activity was measured by Sanofi laboratories using liquid chromatography tandem mass spectrometry (LC-MS/MS) from dried blood spots in a multiplex assay. Dried blood spot punches were incubated with a reaction cocktail containing enzyme specific substrates and buffer. Enzyme activities were quantified by measuring the amount of product generated during incubation. In the PPMI cohort, enzyme activities were measured using a similar method, with addition of a freeze thaw cycle.^18^ In brief, frozen whole blood was slowly thawed on watered ice within 45–60 min. Once thawed, DBS from frozen blood was stored in individual sealed plastic bags with a desiccant at −80°C until analysis. Most of the participants from the PPMI cohort have provided blood samples for three consecutive years. The mean activity across all time points was calculated for the PPMI cohort. Outliers were defined as values exceeding three standard deviations above or below the mean and were excluded from the analysis.

### Statistical analysis

GWAS on the combined cohort was performed using PLINK2^22^ linear regression, with the ratio of glucocerebrosidase to galactosylceramidase as dependent variable, and age, sex, PD status, and first four genomic principal components as covariates (based on scree plot). Sex stratified analyses were performed by conducting GWAS separately on the combined cohort once in females and once in males. Subsequently, GWAS of the glucocerebrosidase/galactosylceramidase ratio was performed separately in the Columbia and PPMI cohorts using PLINK2 linear regression, with age, sex, PD status, and the first four principal components included as covariates. Meta-analysis using summary statistics of Columbia and PPMI cohort was performed using an inverse-variance weighted fixed-effect implemented in METAL using the STDERR scheme.^23^

For selected variants, including the lead SNPs from the genome-wide significant loci identified in the ratio GWAS and the PD GWAS index variant at the *GALC* locus, Pearson correlation coefficients between glucocerebrosidase and galactosylceramidase activities were calculated separately for each genotype group using the cor.test() function in R. Differences between pairs of correlation coefficients were assessed for statistical significance using Fisher’s Z-transformation tests implemented using the R package cocor.^24^

For these selected variants linear regression models of the form galactosylceramidase activity ∼ glucocerebrosidase activity + SNP + glucocerebrosidase activity × SNP + covariates (Age + Sex + PC1-4 + cohort + PD status) were fitted to test for the interaction effects between enzyme activity and genotype. Manhattan plots were generated using R package qqman. Plots of enzymatic activity stratified by genotype were made using the R package ggplot2 {https://ggplot2.tidyverse.org}.

## Results

### Genome-wide association analysis of the glucocerebrosidase/galactosylceramidase ratio

We first performed a GWAS of glucocerebrosidase / galactosylceramidase activity ratio in the combined cohort (N = 1290), adjusting for age, sex, first four principal components and cohort as covariates. The strongest association signal was observed on chromosome 14 at the *GALC* locus (Figure 1A), characterized by a dense set of variants in high linkage disequilibrium. The lead variant at this locus chr14:87945151 (rs365448) showed the strongest association (Effect allele = G, beta = −1.90, P = 5.91 × 10^-115^), which was in LD with the PD GWAS lead variant of the *GALC* locus (rs979812) (D’: 0.628, R^2^: 0.274), which accordingly was also among the variants passing the genome-wide threshold (Effect allele = T, beta = −1.29, P = 1.53 × 10^-46^). Apart from the *GALC* locus, no other loci reached genome-wide significance in the combined-cohort GWAS.

**Figure 1.**
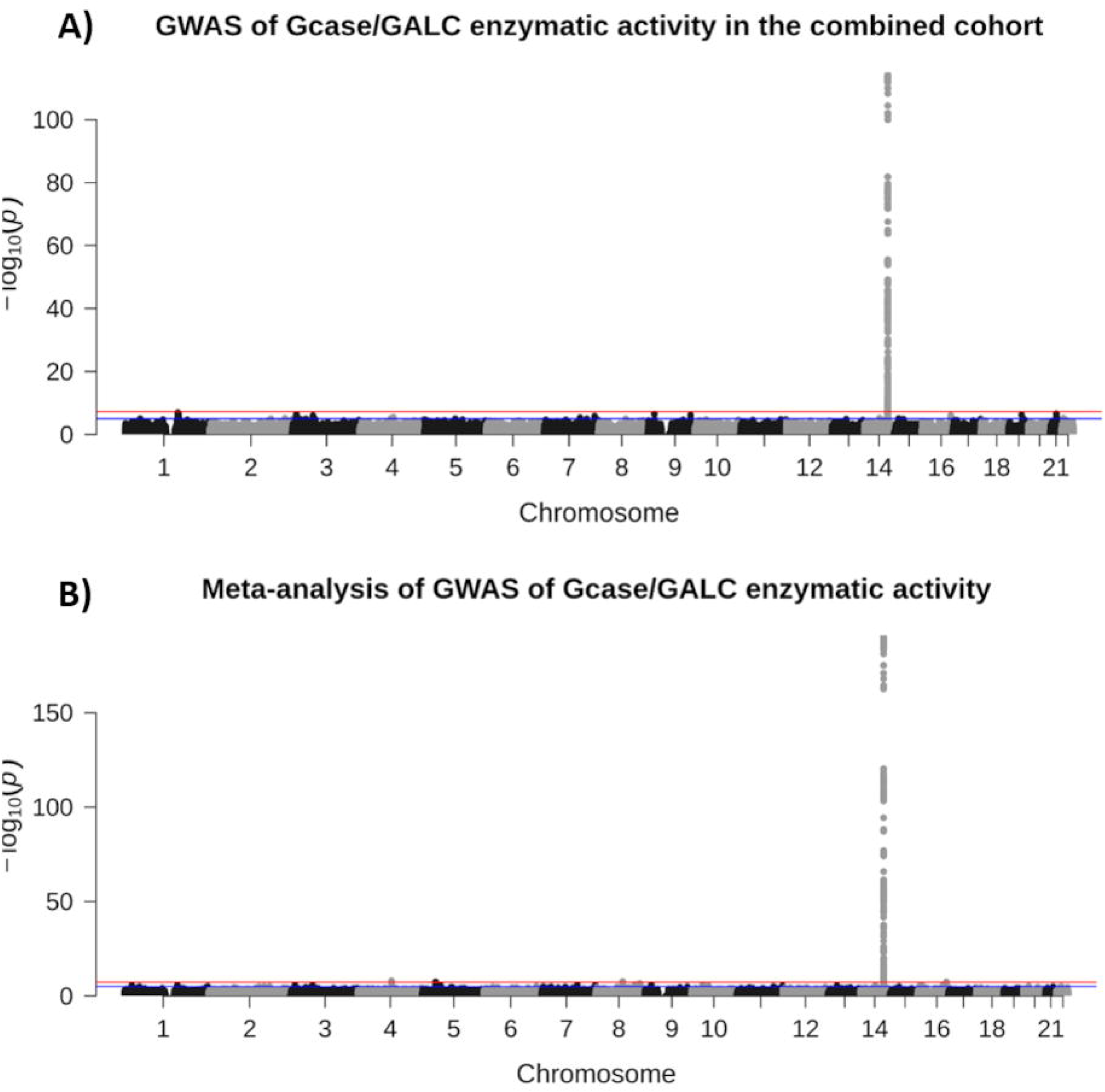
Genome-wide association analyses of the glucocerebrosidase/galactosylceramidase activity ratio. (A) Manhattan plot of the genome-wide association analysis of the glucocerebrosidase/galactosylceramidase activity ratio in the Columbia and PPMI cohort combined. (B) Manhattan plot of the meta-analysis of the combined cohort for glucocerebrosidase/galactosylceramidase activity ratio. The strongest association signal in both analyses was observed at the GALC locus. The x-axis represents chromosomal position, and the y-axis represents -log_10_(P).

To examine whether the findings from the combined cohort analysis are consistent across both cohorts, we analyzed the Columbia and PPMI cohorts separately, followed by a meta-analysis glucocerebrosidase/galactosylceramidase activity ratio (Figure 1B). Both cohorts showed consistent patterns with strong associations in the *GALC* (Figure S1A and S1B), with the same lead variant (rs365448) associated with in the meta-analysis as in the combined cohort (beta = −1.66, P = 6.33 × 10^-191^). Although the direction of effect was consistent across cohorts, the effect size was larger in the Columbia cohort, resulting in substantial between-cohort heterogeneity for this lead variant (I^2^=95.7%, P_het = 1.45 × 10^-6^). Apart from the *GALC* locus, a few additional SNPs reached genome-wide significance in the meta-analysis (Table S1). Quantile-quantile plots showed minimal genomic inflation (Figure S2A and S2B). However, the number of individuals homozygous for the minor allele was too small to allow reliable estimation of correlation coefficients.

### Sex stratified genome-wide association analysis of the glucocerebrosidase / galactosylceramidase activity ratio

A recent sex stratified PD GWAS ^15^ suggested sex-specific effects at the *GALC* locus, with genome-wide significant association observed only in females. Therefore, we also performed our activity ratio GWAS in each sex separately (N_females_ = 522, N_males_ = 768) to determine whether associations with the glucocerebrosidase/galactosylceramidase ratio differed by sex (Figure 2A and 2B).

**Figure 2.**
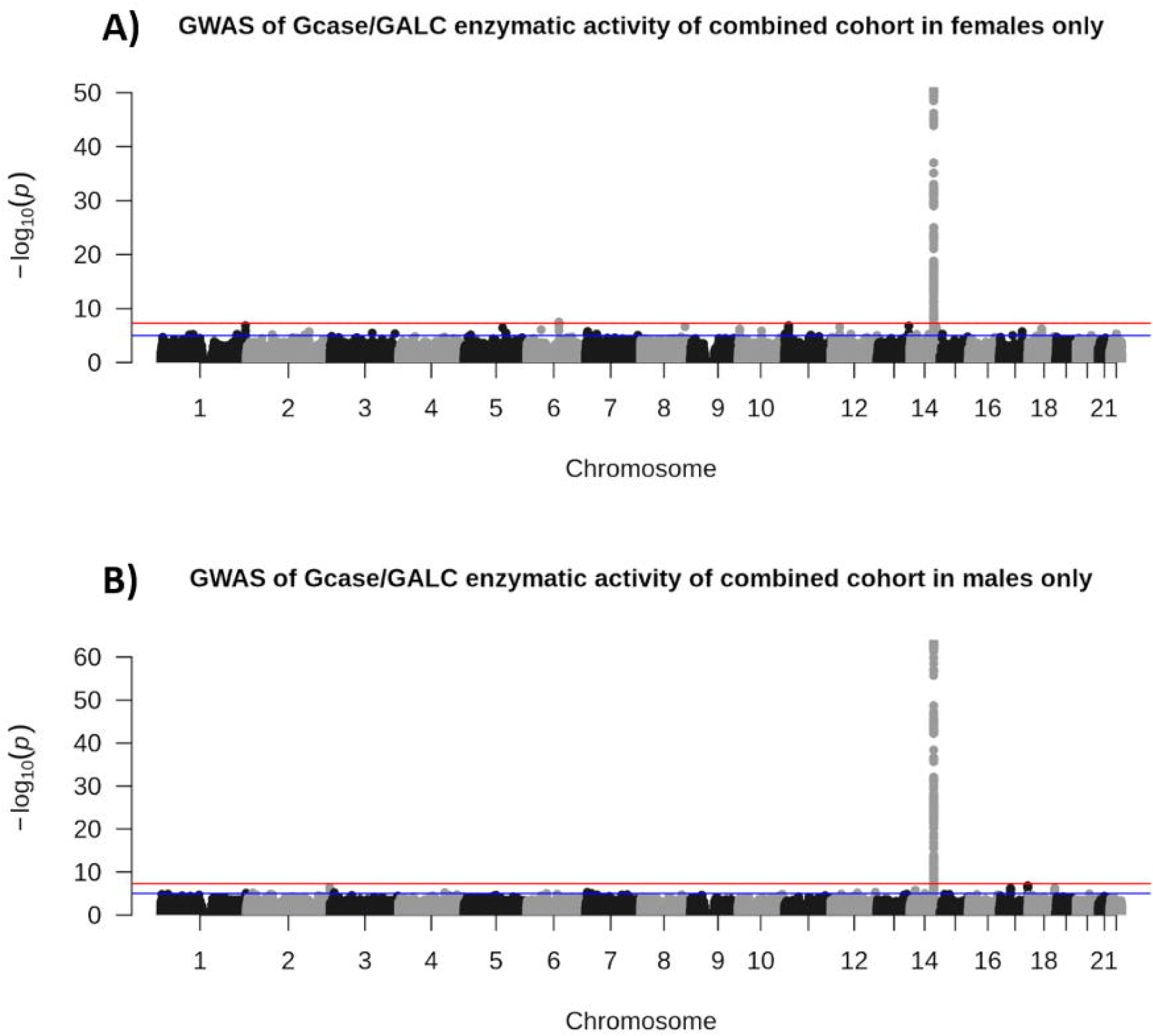
Sex-stratified genome-wide association analyses of the glucocerebrosidase/galactosylceramidase activity ratio in the combined cohort. (A) Manhattan plot of the genome-wide association analysis performed in females in combined cohort. (B) Manhattan plot of the genome-wide association analysis performed in males in combined cohort. The strongest association signal in both analyses was observed at the GALC locus. The x-axis represents chromosomal position, and the y-axis represents -log_10_(P).

Sex-stratified GWAS showed that *GALC* locus remained genome-wide significant in both females and males for the glucocerebrosidase / galactosylceramidase activity ratio. The lead variant for female-GWAS was same as the combined-cohort and meta-analysed cohort (beta = - 1.94, P = 1.60 × 10^-51^) and for male-GWAS was chr14:87938710 (rs424977) (effect allele = A, beta = −1.86, P = 2.43 × 10^-64^) although these variants are in almost perfect linkage disequilibrium with each other (D’: 1.0, R^2^: 0.99). In females, an additional locus near *EPHA7* on chromosome 6 reached genome-wide significance (chr6:93376082, beta = 1.30, P-value = 3.34×10^-8^) that was not detected in the combined analysis. A formal SNP × Sex interaction test for the lead variant in this locus in the combined cohort showed that the effect differed by sex (beta of interaction = 0.615, P-value of interaction = 5.11×10^-5^), although the interaction itself did not reach genome-wide significance. Therefore, this finding should be considered preliminary and requires replication.

### Enzyme activity stratified by genotype at the *GALC* locus

We next examined the relationship between glucocerebrosidase and galactosylceramidase activities at the *GALC* locus, stratified by the PD GWAS index variant rs979812. Interestingly, we found that the relationship between enzymatic activities was different for the different genotypes (Figure 3). Calculating Pearson’s correlation coefficients showed that individuals with TT genotype (T being the effect allele for this variant from the PD GWAS) had a higher correlation (r = 0.33) between enzymatic activities compared to those with the GG genotype (r=0.13), (P = 0.01, Fisher’s Z-transformation test). Stratification by sex showed the same trend in both males and females at the *GALC* locus (Figure S3 and S4). Linear regression with interaction term identified signification interaction between rs979812 genotype and glucocerebrosidase activity, indicating a genotype-dependent association between glucocerebrosidase and galactosylceramidase activities and genotypes (P-value of interaction = 3.05 × 10^-5^).

**Figure 3.**
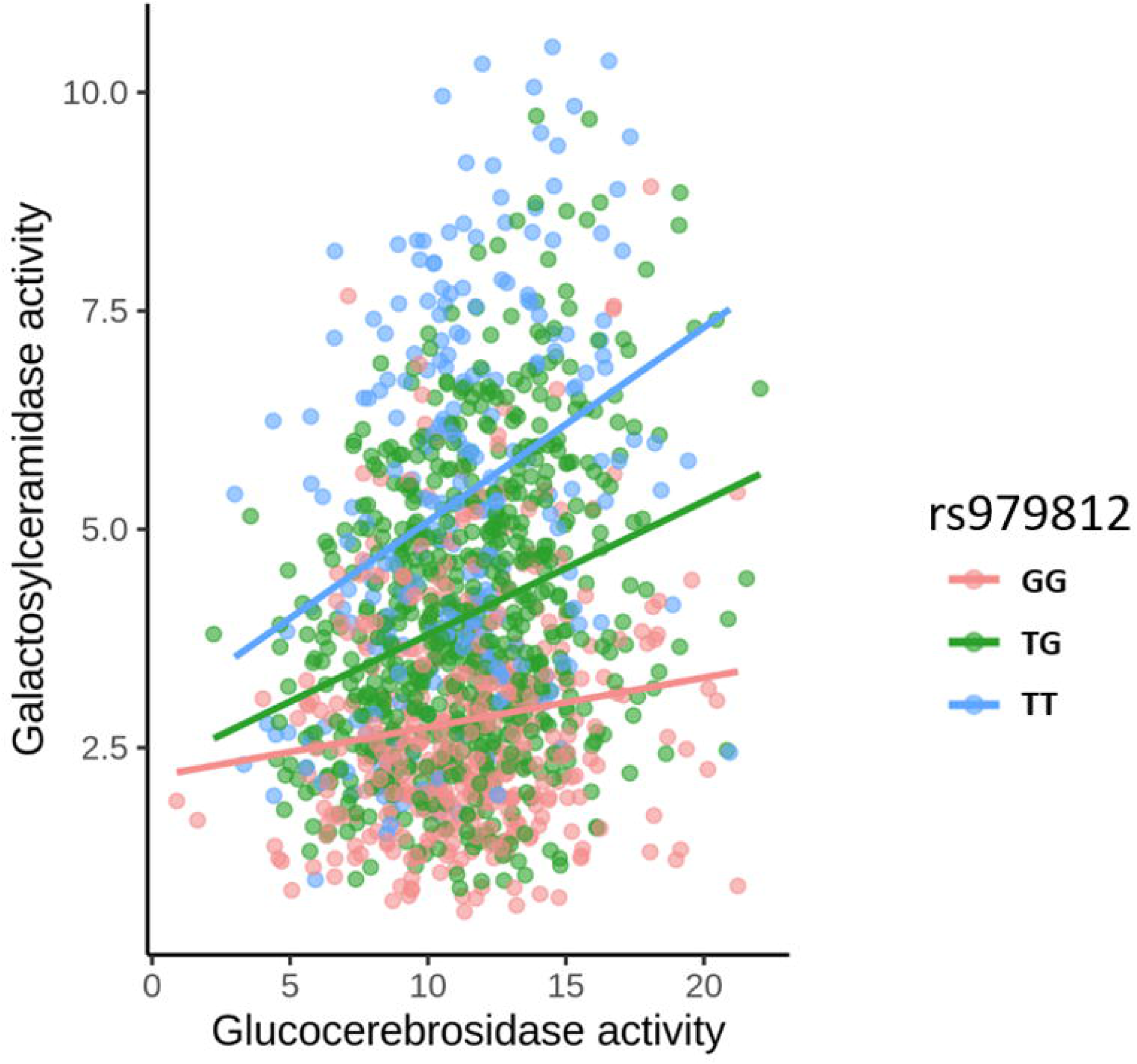
Scatterplot of enzymatic activities of glucocerebrosidase and galactosylceramidase stratified by genotype at the PD GWAS index variant rs979812 (chr14:87997920) at GALC locus in the combined cohort. Each point represents an individual participant, and lines represent genotype-specific linear regression fits. Individuals with GG genotype are shown in red, TG heterozygotes in green, and TT homozygotes in blue. The x-axis represents glucocerebrosidase activity (µmol/l/h), and the y-axis represents galactosylceramidase activity (µmol/l/h).

### Closer look at the PD GWAS lead variants

We further examined specifically the result of the ratio-GWAS from the meta-analysis for the 134 loci from the latest PD GWAS, along with genotype-stratified Pearson correlation coefficients for each genotype for each locus. Ten PD GWAS loci showed nominal associations with the glucocerebrosidase/galactosylceramidase activity ratio apart from the *GALC* locus (Table S2). We calculated the Pearson correlation coefficients between glucocerebrosidase and galactosylceramidase activities in each of these 10 loci, and generated scatterplots (Figure S5 and Table S3). Notably, the *GBA1* locus showed similar trend to *GALC*, with the effect allele carriers showing a higher correlation between the two enzyme activities than non-carriers. Specifically, the PD-associated *GBA1* risk variants p.E326K and p.T369M had higher correlation than non-carriers (Figure 4A and 4B). However, this observation should be interpreted with caution because of the small number of heterozygous carriers, which limits the reliability of the correlation estimate for this locus.

**Figure 4.**
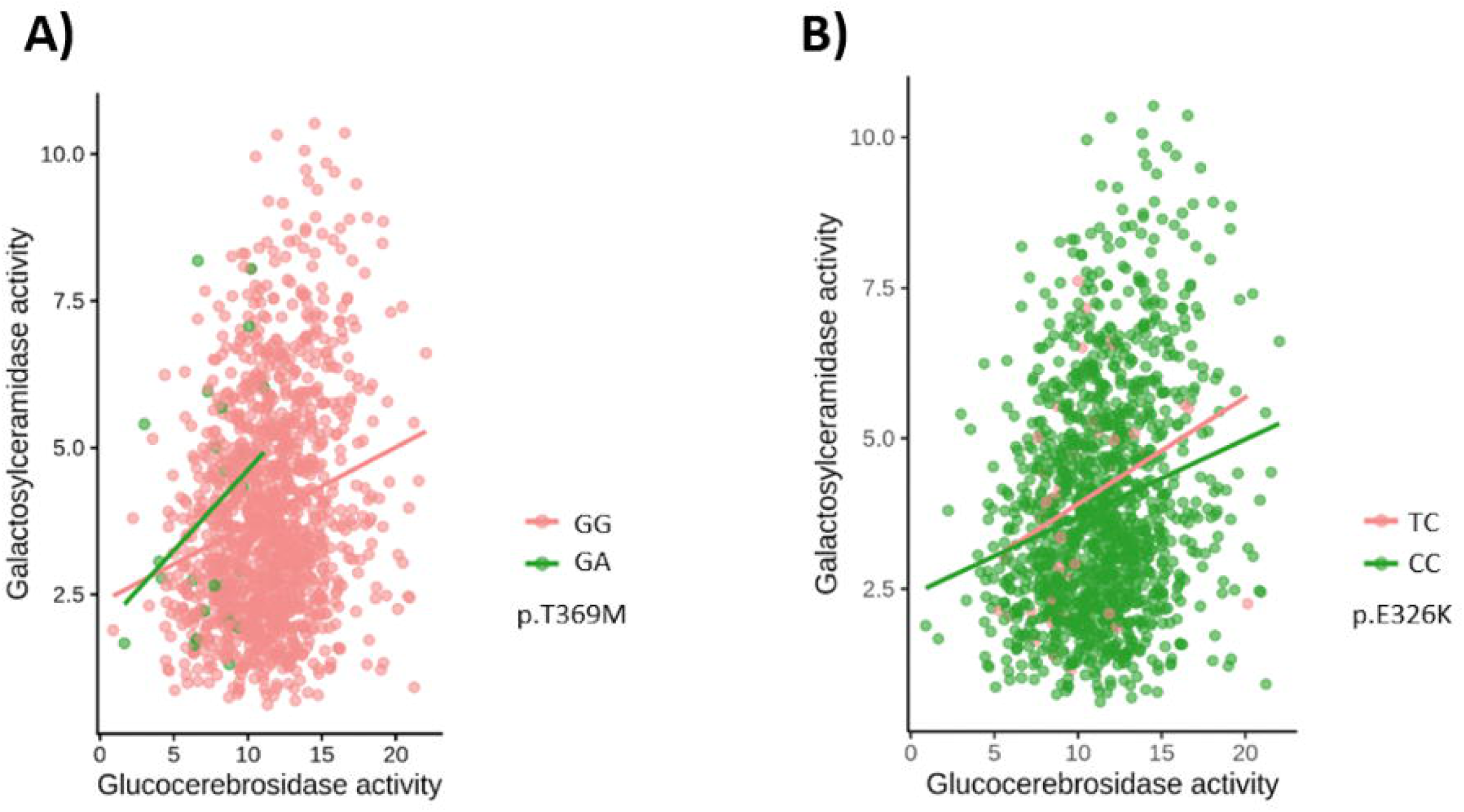
Relationship between glucocerebrosidase and galactosylceramidase activities stratified by GBA1 variant genotype. (A) Scatterplot of glucocerebrosidase and galactosylceramidase activities stratified by GBA1 risk variant p.T369M. GG homozygotes are shown in red and GA heterozygotes are shown in green. (B) Scatterplot of glucocerebrosidase and galactosylceramidase activities stratified by GBA1 risk variant p.E326K. CC homozygotes are shown in green and TC heterozygotes are shown in red. The x-axis represents glucocerebrosidase activity (µmol/l/h), and the y-axis represents galactosylceramidase activity (µmol/l/h).

## Discussion

Our results show a robust, highly significant association between common variants at the *GALC* locus and the glucocerebrosidase-to-galactosylceramidase activity ratio, consistent across two independent cohorts and reinforced by meta-analysis. The lead variant was in linkage disequilibrium with the PD GWAS index variant at this locus, linking *GALC*-associated PD risk to altered enzymatic balance within the lysosomal glycosphingolipid pathway. Notably, the correlation between glucocerebrosidase and galactosylceramidase activities differed significantly by genotype at this locus, with a significant genotype-by-glucocerebrosidase-activity interaction, suggesting that *GALC* variants may influence the coordination between these two enzymes rather than acting on galactosylceramidase alone. A similar, though less robust pattern, was observed at the *GBA1* locus, which requires additional replications. Together, these findings suggest that PD-associated variation at lysosomal glycosphingolipid genes may relate to disease risk partly through effects on the coupled activity of multiple pathway enzymes, rather than through isolated single-enzyme effects.

One possible explanation for these findings is that glucocerebrosidase and galactosylceramidase activities are normally subject to reciprocal, compensatory regulation within the shared lysosomal glycosphingolipid pathway: an increase in flux through one enzyme may trigger a homeostatic adjustment that dampens activity through the other, buffering overall pathway output and limiting substrate accumulation. Such cross-regulation among lysosomal hydrolases has precedent - overexpression of a single glycosphingolipid-degrading enzyme alters the expression of several other lysosomal glycohydrolases, including glucocerebrosidase itself, consistent with a coordinated regulatory network rather than independently controlled enzymes.^25^ This coordination may be mediated, at least in part, by transcriptional programs such as the TFEB/CLEAR network that jointly regulate lysosomal biogenesis and hydrolase expression.^26, 27^ We hypothesize that in carriers of the PD risk-associated *GALC* variant, this compensatory relationship is partially disrupted: rather than glucocerebrosidase and galactosylceramidase activities moving in opposite directions to maintain pathway balance, both trend in the same direction, yielding the stronger positive correlation observed in risk-allele carriers. Under this model, the higher correlation would reflect a loss of independent regulatory buffering rather than a gain of coordinated function, potentially resulting in periods of imbalanced flux through the pathway. This hypothesis will require functional validation in cellular models with defined *GALC* genotypes and isogenic controls.

Another possible hypothesis is that the two activities are linked directly through their shared biochemistry. Glucocerebrosidase and galactosylceramidase act on almost identical substrates. Glucosylceramide and galactosylceramidase differ only in the sugar (glucose vs galactose) on the same ceramide backbone, and both enzymes produce ceramide. *GALC* variants could shift the shared glycosphingolipid or ceramide pool producing a genotype-dependent correlation without any reciprocal regulation of the two enzymes. To examine this hypothesis, one would have to obtain separate substrate measures, not just enzyme activities, which are not currently available.

This study has several limitations. First, our design is cross-sectional and correlational, enzymatic activities were measured at a single time point, so we cannot establish the direction or dynamics of the proposed compensatory relationship, only that it differs by genotype. Second, glucocerebrosidase and galactosylceramidase activities were measured in dried blood spots, a peripheral surrogate that may not fully reflect enzymatic activity or regulation within the central nervous system, where PD-relevant pathology occurs. Furthermore, it was shown that the amount and types of WBC could affect the measured enzymatic activity. The latter may have less of an effect on the results since we examined the ratio between enzymatic activities, where WBC count and type may have the same effect on the nominator and denominator. Third, several findings such as the additional meta-analysis loci, the female-specific *EPHA7* signal, and the *GBA1* correlation pattern, were based on small numbers of variant carriers, limiting the precision of the correlation estimates and requiring cautious interpretation pending replication. Fourth, because we measured only two enzymes, our data cannot distinguish genotype-specific coordinate regulation of glucocerebrosidase and galactosylceramidase from a more general, *GALC*-independent shift in overall lysosomal enzyme capacity between individuals and profiling additional glycosphingolipid-pathway enzymes would help resolve this. Finally, the compensatory-regulation model we propose is a hypothesis motivated by the observed genotype-dependent correlation pattern and existing literature on coordinated lysosomal hydrolase regulation. It was not directly tested here and requires functional validation.

To conclude, we show that common variation at the *GALC* locus is robustly and reproducibly associated with the glucocerebrosidase-to-galactosylceramidase activity ratio, and that the relationship between these two enzyme activities differs by genotype at the PD-associated variant, with a similar, more tentative pattern at *GBA1*. These findings suggest that PD risk at lysosomal glycosphingolipid genes may involve altered coordination between enzymes within the pathway, in addition to effects on any single enzyme. Future work should test this directly using patient-derived cellular models with defined *GALC* and *GBA1* genotypes and isogenic controls to determine whether manipulating one enzyme’s activity produces the expected compensatory, genotype-dependent response in the other, and whether this coupling is disrupted in a directional, mechanistically interpretable way. Extending the analysis to longitudinal follow-up data would help clarify whether this activity coupling reflects a stable trait or one that changes with disease progression, and whether it holds diagnostic or pharmacodynamic value. Given ongoing efforts to develop GCase-targeted therapies for *GBA1*-associated PD, it will also be worth testing whether such interventions produce off-target or compensatory shifts in galactosylceramidase activity, which could serve as a pathway-level marker of target engagement.

## Acknowledgements

We thank the research participants for contributing to this study. This study was financially supported through grants from the Michael J. Fox Foundation. This paper was supported by the Galen and Hilary Weston Foundation and received contributions from the G-Can (GBA1-Canada) Initiative. G-Can is supported by The Hilary and Galen Weston Foundation, Silverstein Foundation, and J. Sebastian van Berkom and Ghislaine Saucier.

Data used in the preparation of this article were obtained from the Parkinson’s Precision Medicine Initiative (PPMI) database (www.ppmi-info.org/access-data-specimens/download-data), RRID:SCR_006431. For up-to-date information on the study, visit www.ppmi-info.org. The Columbia University cohort is supported by the Parkinson’s Foundation, the National Institutes of Health (K02NS080915, and UL1 TR000040) and the Brookdale Foundation.

Computational analyses performed in this study were supported by NeuroHub, Calcul Québec, and the Digital Research Alliance of Canada.

Z.G-O. is supported by the Fonds de recherche du Québec—Santé (FRQS) Chercheurs-boursiers award and is a William Dawson Scholar. K.S. is supported by Parkinson Canada. A.H. is supported by a Clinical Research Scholarship (349722) and a Clinician-Researcher Establishment Grant (358114) from the Fonds de recherche du Québec—Santé (FRQS), the Bougie Family Young Investigator Award, the Canadian Institutes of Health Research (CIHR), and a Future Leaders in Canadian Brain Research Grant from the Brain Canada Foundation. E.F is supported by a Canada Research Chair (Tier 1) in Parkinson’s disease.

## Author contributions

A.R.V, K.S, and Z.G.O contributed to the conception and design of the study. All authors contributed to the acquisition and analysis of the data. A.R.V and Z.G.O contributed to drafting a significant portion of the manuscript or figures.

## Potential conflicts of Interest

Z.G.O. received consultancy fees from Lysosomal Therapeutics (LTI), Idorsia, Prevail Therapeutics, Ono Therapeutics, Denali, Handl Therapeutics, Neuron23, Bial Biotech, Bial, UCB, Capsida, Vanqua bio, Simcere, Jazz therapeutics, Takeda, EG427, Gain Therapeutics, Guidepoint, Lighthouse, and Deerfield. K.S. received consultancy fees from Acurex. A.H. has received speaker and/or consultancy fees from Alexion, Amgen, Biogen, Novartis, Pfizer, and Roche; compensation for serving on a scientific advisory board for Amgen, Novartis, and Roche; and research funding from Amgen and Roche, all unrelated to the present work. Other authors declare no conflicts of interest.

## Data Availability

Data used in the preparation of this article were obtained from the Parkinson’s Progression Markers Initiative (PPMI) database (www.ppmi-info.org/access-data-specimens/download-data). For up-to-date information on the study, visit www.ppmi-info.org. Columbia cohort data can be obtained by request. The ratio-GWAS summary statistics can be found on the GWAS catalog (https://www.ebi.ac.uk/gwas/) accession number (GCST91000565, GCST91000566, GCST91000567, GCST91000568, GCST91000569, GCST91000570). Variants that pass genome-wide significance for each GWAS can be found in the supplementary tables S4-S9. All code for analyses used in this project can be found at gan-orlab GitHub page. PPMI data used for this study can be obtained by qualified researchers upon completion of a data access application (https://www.ppmi-info.org/access-data-specimens/download-data).

## Notes

### Author Declarations

McGill University Research Ethics Board Office gave ethical approval (21-11-023).

## References

1. von Linstow CU, Gan-Or Z, Brundin P. Precision medicine in Parkinson’s disease patients with LRRK2 and GBA risk variants - Let’s get even more personal. Transl Neurodegener. 2020 Oct 16;9(1):39.

2. Leonard HL, Global Parkinson’s Genetics P. Novel Parkinson’s Disease Genetic Risk Factors Within and Across European Populations. medRxiv. 2025 Mar 17.

3. Lange LM, Cerquera-Cleves C, Schipper M, et al. Prioritizing Parkinson’s disease risk genes in genome-wide association loci. NPJ Parkinsons Dis. 2025 Apr 16;11(1):77.

4. Yu E, Lariviere R, Thomas RA, et al. Machine learning nominates the inositol pathway and novel genes in Parkinson’s disease. Brain. 2024 Mar 1;147(3):887–99.

5. Sidransky E, Nalls MA, Aasly JO, et al. Multicenter analysis of glucocerebrosidase mutations in Parkinson’s disease. N Engl J Med. 2009 Oct 22;361(17):1651–61.

6. Senkevich K, Zorca CE, Dworkind A, et al. GALC variants affect galactosylceramidase enzymatic activity and risk of Parkinson’s disease. Brain. 2023 May 2;146(5):1859–72.

7. Alcalay RN, Mallett V, Vanderperre B, et al. SMPD1 mutations, activity, and alpha-synuclein accumulation in Parkinson’s disease. Mov Disord. 2019 Apr;34(4):526–35.

8. Robak LA, Jansen IE, van Rooij J, et al. Excessive burden of lysosomal storage disorder gene variants in Parkinson’s disease. Brain. 2017 Dec 1;140(12):3191–203.

9. Senkevich K, Gan-Or Z. Autophagy lysosomal pathway dysfunction in Parkinson’s disease; evidence from human genetics. Parkinsonism Relat Disord. 2020 Apr;73:60–71.

10. Gan-Or Z, Liong C, Alcalay RN. GBA-Associated Parkinson’s Disease and Other Synucleinopathies. Curr Neurol Neurosci Rep. 2018 Jun 8;18(8):44.

11. Rizig M, Bandres-Ciga S, Makarious MB, et al. Identification of genetic risk loci and causal insights associated with Parkinson’s disease in African and African admixed populations: a genome-wide association study. Lancet Neurol. 2023 Nov;22(11):1015–25.

12. Alvarez Jerez P, Wild Crea P, Ramos DM, et al. African ancestry neurodegeneration risk variant disrupts an intronic branchpoint in GBA1. Nat Struct Mol Biol. 2024 Dec;31(12):1955–63.

13. Boer DEC, van Smeden J, Bouwstra JA, Aerts J. Glucocerebrosidase: Functions in and Beyond the Lysosome. J Clin Med. 2020 Mar 9;9(3).

14. Bongarzone ER, Escolar ML, Gray SJ, Kafri T, Vite CH, Sands MS. Insights into the Pathogenesis and Treatment of Krabbe Disease. Pediatr Endocrinol Rev. 2016 Jun;13 Suppl 1:689–96.

15. Global Parkinson’s Genetics P, Leonard HL. Shared and Sex-Specific Genetic Risk for Parkinson’s Disease Risk Across European Populations. medRxiv. 2026 Jan 2.

16. Marek K, Chowdhury S, Siderowf A, et al. The Parkinson’s progression markers initiative (PPMI) - establishing a PD biomarker cohort. Ann Clin Transl Neurol. 2018 Dec;5(12):1460–77.

17. Alcalay RN, Levy OA, Waters CC, et al. Glucocerebrosidase activity in Parkinson’s disease with and without GBA mutations. Brain. 2015 Sep;138(Pt 9):2648–58.

18. Alcalay RN, Wolf P, Chiang MSR, et al. Longitudinal Measurements of Glucocerebrosidase activity in Parkinson’s patients. Ann Clin Transl Neurol. 2020 Oct;7(10):1816–30.

19. Purcell S, Neale B, Todd-Brown K, et al. PLINK: a tool set for whole-genome association and population-based linkage analyses. Am J Hum Genet. 2007 Sep;81(3):559–75.

20. Reuser AJ, Verheijen FW, Bali D, et al. The use of dried blood spot samples in the diagnosis of lysosomal storage disorders--current status and perspectives. Mol Genet Metab. 2011 Sep-Oct;104(1-2):144–8.

21. Zhang XK, Elbin CS, Chuang WL, et al. Multiplex enzyme assay screening of dried blood spots for lysosomal storage disorders by using tandem mass spectrometry. Clin Chem. 2008 Oct;54(10):1725–8.

22. Chang CC, Chow CC, Tellier LC, Vattikuti S, Purcell SM, Lee JJ. Second-generation PLINK: rising to the challenge of larger and richer datasets. Gigascience. 2015;4:7.

23. Willer CJ, Li Y, Abecasis GR. METAL: fast and efficient meta-analysis of genomewide association scans. Bioinformatics. 2010 Sep 1;26(17):2190–1.

24. Diedenhofen B, Musch J. cocor: a comprehensive solution for the statistical comparison of correlations. PLoS One. 2015;10(3):e0121945.

25. Tancini B, Magini A, Bortot B, et al. Beta-hexosaminidase over-expression affects lysosomal glycohydrolases expression and glycosphingolipid metabolism in mammalian cells. Mol Cell Biochem. 2012 Apr;363(1-2):109–18.

26. Sardiello M, Palmieri M, di Ronza A, et al. A gene network regulating lysosomal biogenesis and function. Science. 2009 Jul 24;325(5939):473–7.

27. Napolitano G, Ballabio A. TFEB at a glance. J Cell Sci. 2016 Jul 1;129(13):2475–81.

