## Supplementary Figures for "*GALC* variants affect glucocerebrosidase to galactosylceramidase activity ratio"

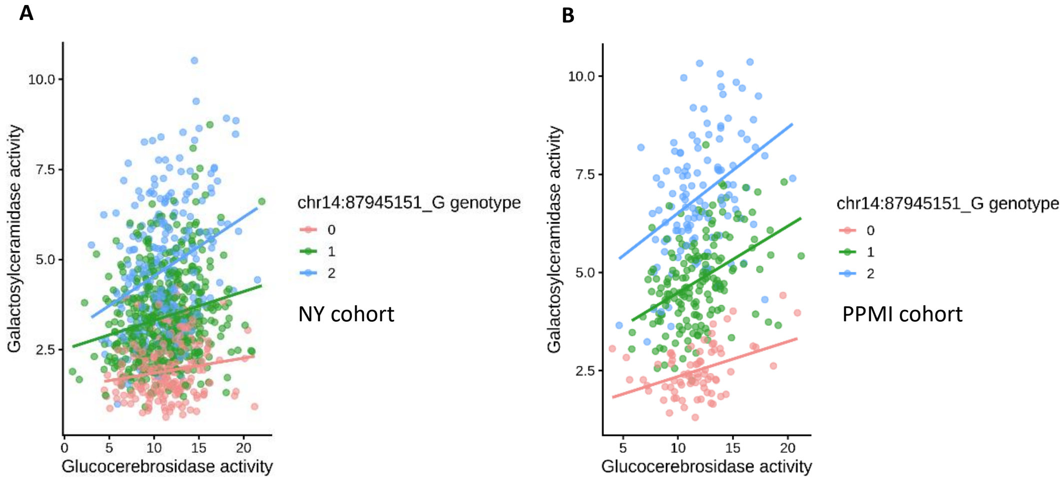


***Figure S1. Relationship between glucocerebrosidase and galactosylceramidase activities stratified by rs365448 genotype in each individual cohort.*** *(A) Scatterplot of glucocerebrosidase and galactosylceramidase activities in the Columbia cohort stratified by rs365448 genotype. (B) Corresponding analysis in the PPMI cohort. Each point represents an individual participant, and solid lines represent genotype-specific linear regression fits. Genotype groups are coded according to G allele dosage: zero copies are represented as red, one copy in green, and two copies in blue. The x-axis represents glucocerebrosidase activity (µmol/l/h), and the y-axis represents galactosylceramidase activity (µmol/l/h).*

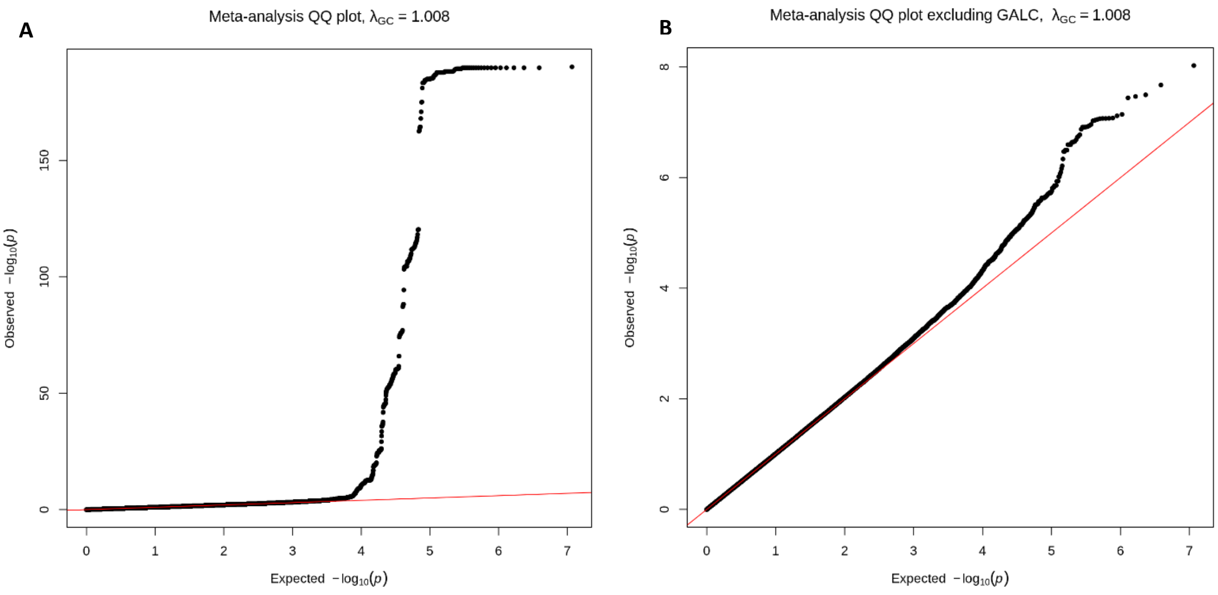


***Figure S2. Assessment of genomic inflation in meta-analysis.*** *(A) Quantile-quantile plot of full meta-analysis. (B) Quantile-quantile plot when the GALC locus is excluded. The x-axis represents the expected -$log_{10}$(P) values from the association analysis and the y-axis represents the observed -$log_{10}$(P) values from the association analysis. The genomic inflation factor was 1.008.*

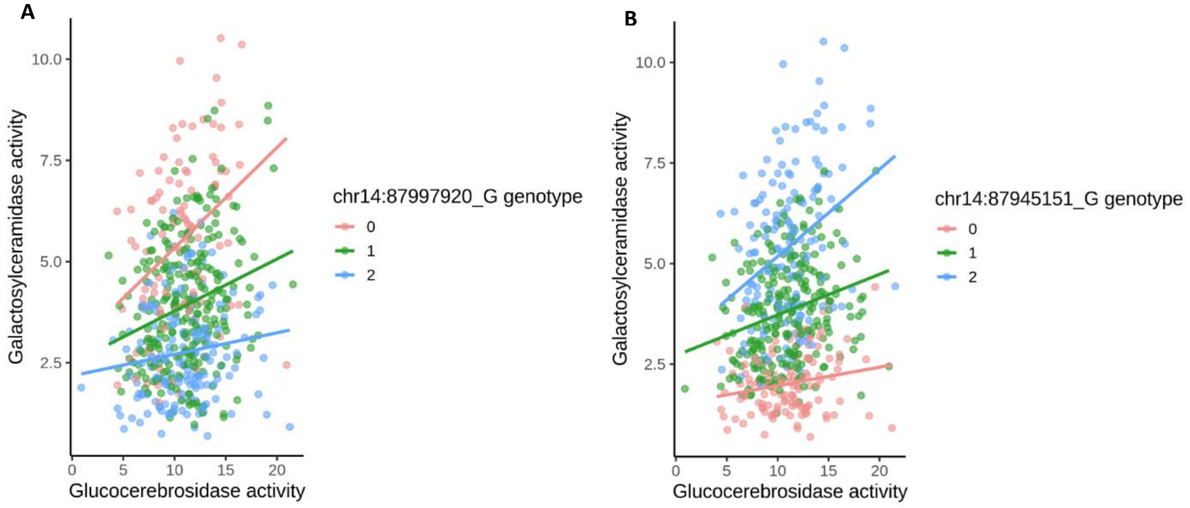


***Figure S3. Sex stratified relationship between glucocerebrosidase and galactosylceramidase activities at the GALC locus in females.*** *(A) Scatterplot of the glucocerebrosidase and galactosylceramidase activities stratified by PD GWAS index variant at the GALC locus (rs979812). (B) Scatterplot of the glucocerebrosidase and galactosylceramidase activities stratified by ratio-GWAS top variant. Each point represents an individual participant, and solid lines represent genotype-specific linear regression fits. Genotype groups are coded according to G allele dosage: zero copies are represented as red, one copy in green, and two copies in blue. The x-axis represents glucocerebrosidase activity (µmol/l/h), and the y-axis represents galactosylceramidase activity (µmol/l/h).*

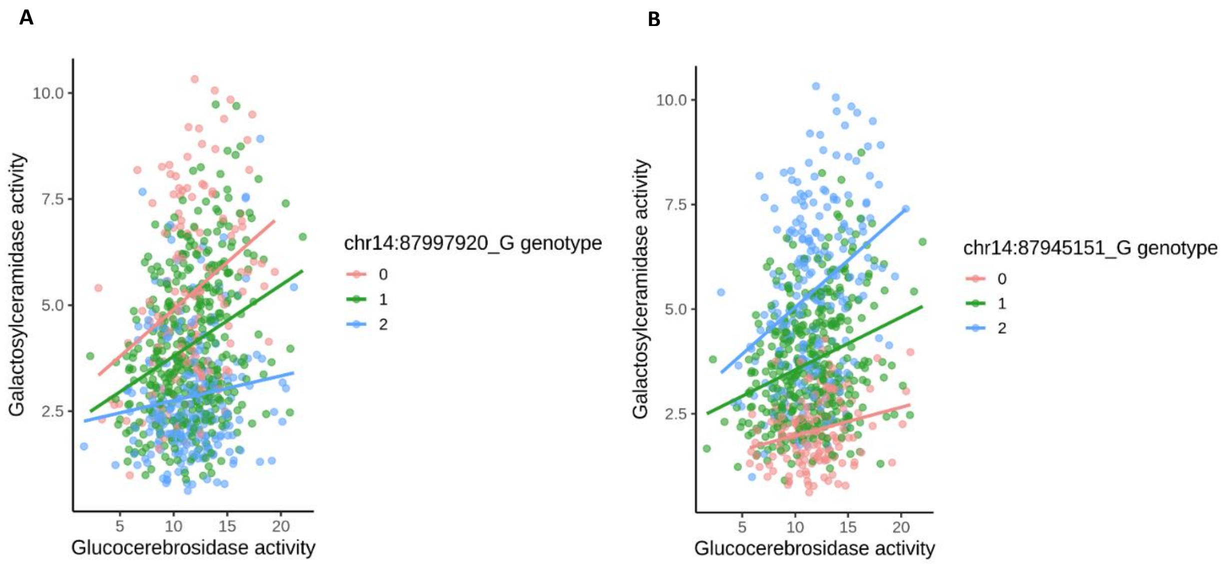


***Figure S4. Sex stratified relationship between glucocerebrosidase and galactosylceramidase activities at the GALC locus in***

***males.*** *(A) Scatterplot of the glucocerebrosidase and galactosylceramidase activities stratified by PD GWAS index variant at the GALC locus (rs979812). (B) Scatterplot of the glucocerebrosidase and galactosylceramidase activities stratified by ratio-GWAS top variant. Each point represents an individual participant, and solid lines represent genotype-specific linear regression fits. Genotype groups are coded according to G allele dosage: zero copies are represented as red, one copy in green, and two copies in blue. The x-axis represents glucocerebrosidase activity (µmol/l/h), and the y-axis represents galactosylceramidase activity (µmol/l/h).*

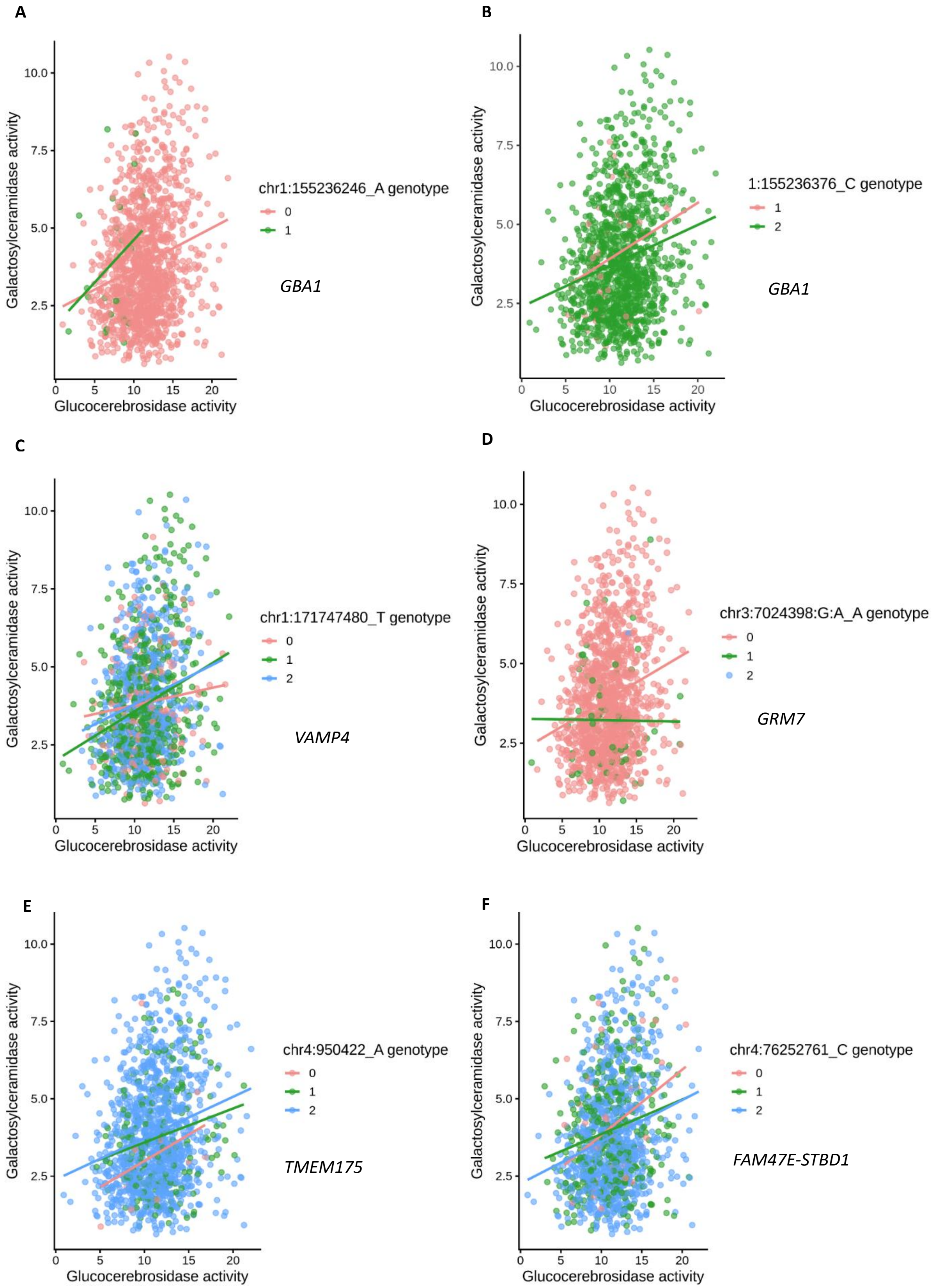
A
Galactosylceramidase activity
Glucocerebrosidase activity
chr1:155236246_A genotype
0
1
GBA1
B
Galactosylceramidase activity
Glucocerebrosidase activity
1:155236376_C genotype
1
2
GBA1
C
Galactosylceramidase activity
Glucocerebrosidase activity
chr1:171747480_T genotype
0
1
2
VAMP4
D
Galactosylceramidase activity
Glucocerebrosidase activity
chr3:7024398:G:A_A genotype
0
1
2
GRM7
E
Galactosylceramidase activity
Glucocerebrosidase activity
chr4:950422_A genotype
0
1
2
TMEM175
F
Galactosylceramidase activity
Glucocerebrosidase activity
chr4:76252761_C genotype
0
1
2
FAM47E-STBD1

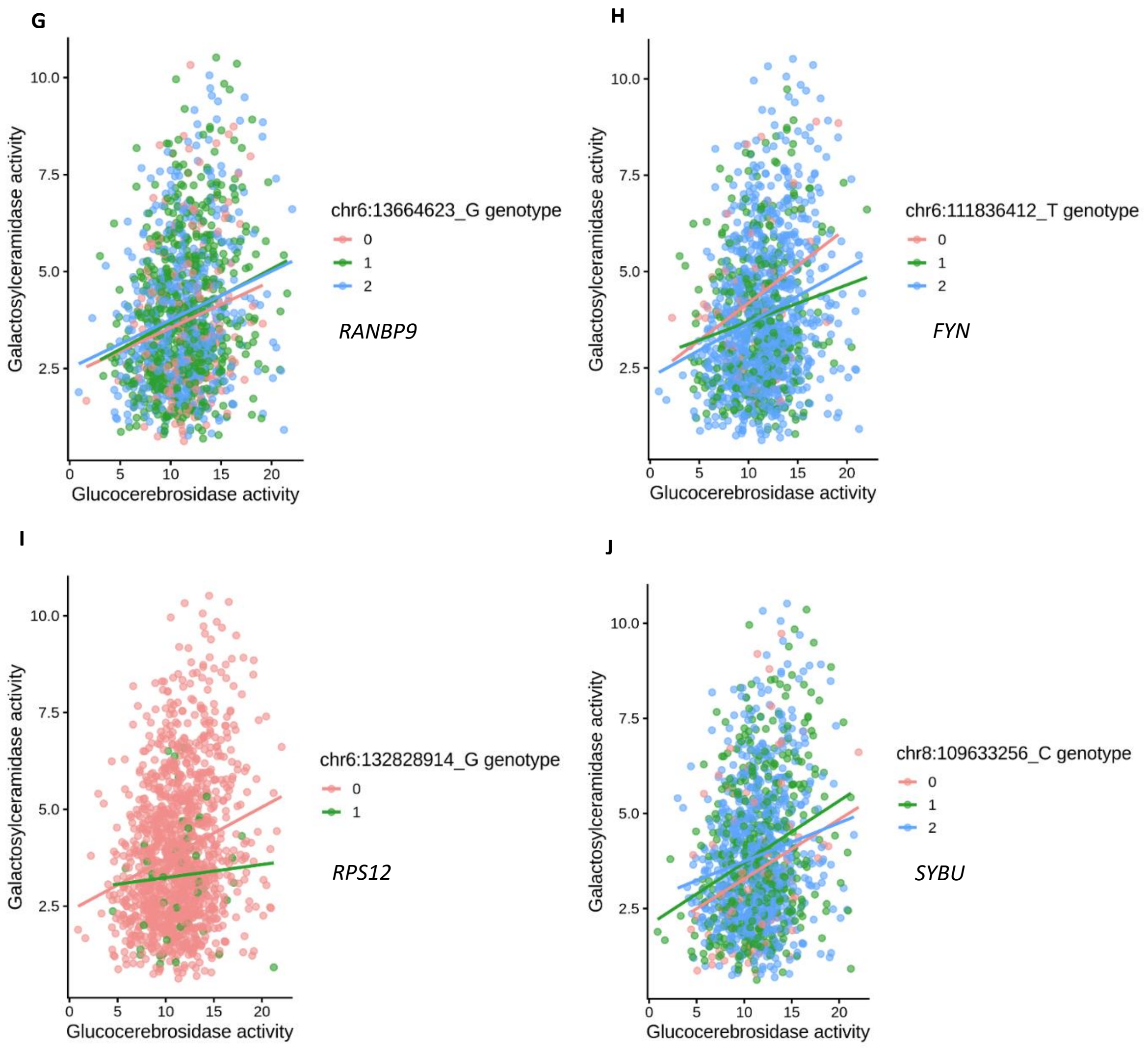
G
Galactosylceramidase activity
Glucocerebrosidase activity
chr6:13664623_G genotype
0
1
2
RANBP9
H
Galactosylceramidase activity
Glucocerebrosidase activity
chr6:111836412_T genotype
0
1
2
FYN
I
Galactosylceramidase activity
Glucocerebrosidase activity
chr6:132828914_G genotype
0
1
RPS12
J
Galactosylceramidase activity
Glucocerebrosidase activity
chr8:109633256_C genotype
0
1
2
SYBU

K

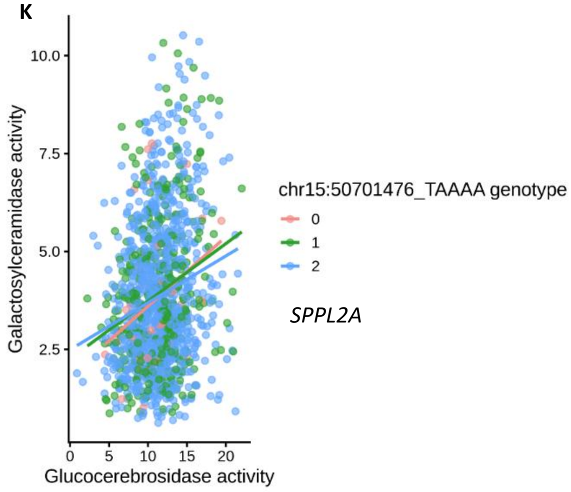


*SPPL2A*

***Figure S5. Scatterplot of PD GWAS lead variants nominally associated in the ratio-GWAS meta-analysis.*** *Panels A-K show the individual lead variant and the nearest gene to it in each panel. Each point represents an individual participant, and solid lines represent genotype-specific linear regression fits. Genotype groups are coded according to the dosage of the allele indicated on each panel: zero copies are represented as red, one copy in green, and two copies in blue. The x-axis represents glucocerebrosidase activity (µmol/l/h), and the y-axis represents galactosylceramidase activity (µmol/l/h).*
